# HELIOS: A spatial omics-informed multi-organ cell atlas enables AI-based tumour microenvironment profiling and biomarker discovery from routine histopathology

**DOI:** 10.64898/2026.10.26.26366087

**Authors:** Zhuoyan Shen, Huiyan Hu, Konstantin Bräutigam, Jinyi Wang, Manuel Rodriguez-Justo, Maria A. Hawkins, Charles-Antoine Collins Fekete

## Abstract

Spatial omics defines tumour microenvironments with high biological resolution but remains difficult to scale across large clinical cohorts. We developed HELIOS (H&E Learning Informed by Omics and Spatial Profiling), which transfers spatial-omics-derived cell identities into routine haematoxylin and eosin histology for interpretable single-cell tumour microenvironment profiling. HELIOS integrates 12.9 million cells from 60 specimens across 13 tissue types and four spatial profiling platforms, spanning eight epithelial, immune, stromal, and vascular populations. The resulting multiclass detector generalised across independent histopathology benchmarks. Applied to colorectal, breast, and lung cancers, HELIOS recapitulated molecular subtype-associated microenvironment states and identified prognostic immune and stromal phenotypes. Tumour-infiltrating lymphocytes were consistently associated with favourable outcomes, whereas neutrophil and fibroblast phenotypes showed cancer-specific associations. HELIOS-derived features remained prognostic after adjustment for clinicopathological and molecular variables. These findings demonstrate that spatial omics can provide scalable biological supervision for extracting clinically relevant tumour microenvironment features from routine H&E, supporting scalable risk stratification and biomarker development across clinical cohorts and retrospective pathology archives.

## Introduction

Histopathological assessment of haematoxylin and eosin (H&E)-stained tissue sections remains central to disease diagnosis, grading, staging, and treatment decision-making. Because H&E slides are generated routinely, they represent a clinically embedded resource for studying tumour morphology and the tumour microenvironment. Digital pathology has enabled AI-based analysis of these specimens at scale, supporting tumour detection, tissue classification, molecular prediction, and prognostic stratification^1–3^. More recently, pathology foundation models have provided a powerful framework for learning transferable visual representations from large histology datasets^4–6^. Although these models have improved performance across multiple downstream tasks, their ability to resolve biologically defined cell types remains limited by the quality and scale of available supervision. As a result, AI-derived histology biomarkers may achieve predictive performance while remaining difficult to interpret mechanistically, limiting their translational value.

Cell-level analysis of the tumour microenvironment is clinically important because the abundance and spatial organisation of tumour, immune, and stromal cells shape cancer progression and therapeutic response^7,8^. Precision oncology has transformed cancer management through molecular biomarkers that guide targeted therapies, such as KRAS, BRAF and EGFR alterations and mismatch repair deficiency/microsatellite instability (dMMR/MSI) status in colorectal cancer and HER2 amplification in breast cancer^9–11^. However, many patients lack actionable molecular alterations or fall within broad, biologically heterogeneous groups, such as microsatellite-stable (MSS) colorectal cancer and HER2-negative breast cancer, for which additional approaches to treatment selection and prognostic stratification are needed^12^. Moreover, substantial variation in clinical outcomes and therapeutic responses persists even among patients within the same molecular subtype^12^. Characterisation of the tumour microenvironment can therefore provide complementary information beyond genomics^13,14^. Features such as tumour-infiltrating lymphocytes, tumour-associated macrophages and tumour-stromal architecture have been linked to prognosis and treatment sensitivity across cancer types^15–17^.

However, accurate nuclei detection and classification, and quantitative assessment of cellular abundance and spatial organisation from routine H&E remains challenging across diverse organs, tumour types, and histological contexts. Existing datasets often rely on manual labels derived from morphology alone and are often restricted to a limited range of tissues and cell types^18–19^. Widely used models such as HoVer-Net trained on Lizard^20^, have advanced this field, but the performance are constrained by the quality and scope of manual annotations, which is not only labour-intensive but also limited by inter-observer variability. Co-registration of H&E with immunohistochemistry or multiplex immunofluorescence has produced more reliable ground truth labels and improved immune-cell classification^21,22^, but these approaches usually assay selected markers, require additional tissue processing, and do not comprehensively capture the cellular complexity of the tumour microenvironment.

The development of high-resolution spatial transcriptomics provides an opportunity to link molecular cell identity with preserved tissue architecture^23,24^. In cancer, these data offer the opportunity to characterise the actual tumour ecosystem and give insight into tumour biology beyond genomics. Direct prediction of spatial gene expression from H&E, sometimes termed virtual spatial transcriptomics, is promising^25,26^ but challenging because transcriptomic states are not always fully encoded in morphology. Rather than treating gene expression as the prediction target, we instead use single-cell spatial transcriptomics as a high-fidelity supervision: gene profiles define cell identities in the matched H&E for training AI models to recover the corresponding cell classes from routine histology.

Here, we present HELIOS, H&E Learning Informed by Omics and Spatial Profiling, a cancer-focused, biology-grounded atlas for AI-based nuclei detection and classification in routine pathology. HELIOS links high-resolution spatial transcriptomics with matched H&E morphology across 60 samples, 13 human tissue types, and eight cell types spanning the major epithelial, immune, and stromal/vascular compartments, including epithelial cells, lymphocytes, fibroblasts, macrophages, vascular-associated cells (endothelial cells and pericytes), plasma cells, smooth-muscle cells, and neutrophils. This taxonomy was chosen to cover recurrent, biologically meaningful tumour microenvironment cell types while being visually learnable from H&E. We then use HELIOS to train an object detection model that detects and classifies cells directly from H&E images. The model generates whole-slide cellular maps and quantitative measures of tumour-microenvironment composition. Across independent cohorts, HELIOS-derived measures of densities of tumour-infiltrating lymphocytes, tumour-associated macrophages, neutrophils and fibroblast-to-epithelial ratio stratify survival in independent cohorts across lung, breast, and colorectal cancers. These AI-derived biomarkers also provided prognostic information beyond conventional tumour grading and established molecular subgroups. To support translation, HELIOS has also been deployed through a web-based platform that is undergoing evaluation in clinical settings. Since it outputs visually inspectable cell maps and interpretable cellular biomarkers, and requires only routine H&E images, HELIOS has the potential to support scalable tumour-microenvironment profiling and phenotyping, and integration into existing digital-pathology workflows.

## Results

### HELIOS: A large-scale multi-organ cell atlas for digital pathology

Figure 1 summarises the HELIOS construction pipeline and data composition. We developed HELIOS by integrating spatial transcriptomics with co-registered H&E images, enabling molecularly defined cell identities to be mapped onto histological morphology at single-cell resolution, followed by quality control and benchmarking against pathologist-derived manual annotations (Figure 1A). Annotation accuracy increased progressively with label confidence and exceeded 0.85 among cells with confidence scores greater than 0.5. Notably, 90% of all annotated cells met this high-confidence threshold, supporting the overall reliability of the resulting cell atlas.

**Figure 1.**
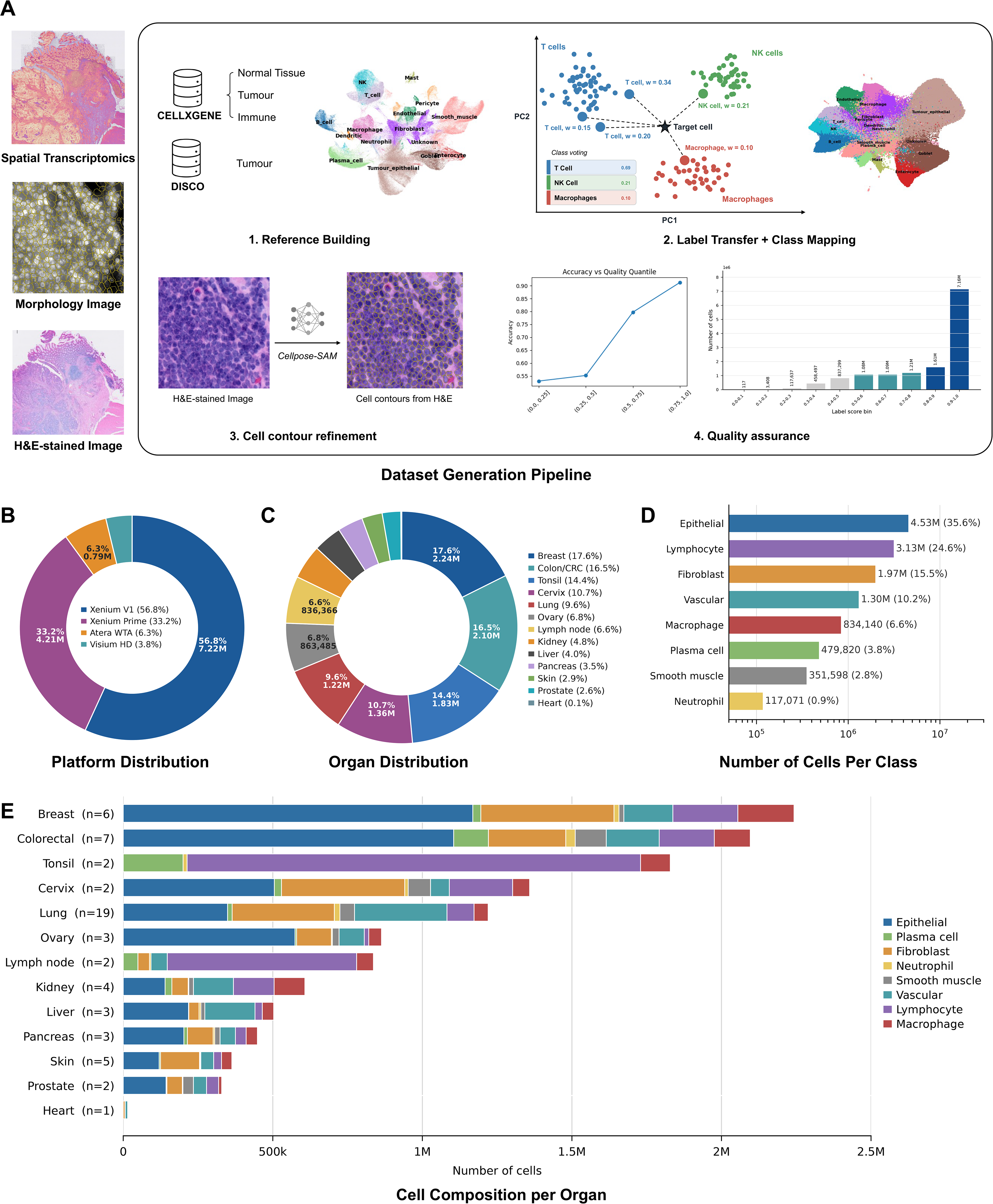
Construction and composition of the HELIOS multi-organ cell atlas. **(A)** Overview of the HELIOS annotation workflow. Spatial transcriptomic profiles were integrated with co-registered H&E images and segmented cell contours. Cell identities were assigned using organ- and gene-panel-matched single-cell RNA-sequencing references, PCA-based nearest-neighbour label transfer, and distance-weighted class scores. Transcriptomic labels were projected onto H&E-derived cell contours and filtered using confidence-based quality control. Manual review showed progressively greater annotation accuracy across increasing confidence-score quantiles; the accompanying histogram shows the distribution of confidence scores across the complete atlas. **(B)** Distribution of annotated cells across the four spatial profiling platforms: Xenium V1, Xenium Prime, Atera WTA, and Visium HD. **(C)** Distribution of annotated cells across 13 human tissue types. **(D)** Number and proportion of cells assigned to each of the eight final HELIOS classes. **(E)** Number and proportion of cells within each organ; values in parentheses indicate the number of spatial transcriptomic samples.

HELIOS integrates independent spatial omics datasets generated across four platforms, including Xenium V1, Xenium Prime, Visium HD, and Atera WTA (Figure 1B). The atlas comprises primary cancer specimens across breast, colorectal, cervical, lung, ovarian, kidney, pancreatic, prostate, liver and skin tissues. To capture the broad morphological diversity encountered in routine histopathology and improve the generalisability of the model, we also include non-malignant tissues, including tonsillar hyperplasia, normal colorectal tissue, normal (prelaryngeal) lymph node, normal kidney tissue, normal liver tissue, normal skin tissue, and normal heart tissue (Figure 1C).

HELIOS contains 12.9 million annotated cells assigned to eight major epithelial, immune, stromal, and vascular populations: epithelial cells, lymphocytes, fibroblasts, macrophages, vascular-associated cells, plasma cells, smooth-muscle cells, and neutrophils (Figure 1D). Cell-type composition varied substantially across organs, reflecting differences in tissue architecture and the organisation of immune, stromal, and vascular compartments (Figure 1E).

### HELIOS supports accurate cell detection and classification across organs

To evaluate whether HELIOS could support accurate cell detection and classification directly from routine H&E images, we trained a multi-class YOLOv10-m model^27^ and assessed performance using precision, recall, and F1 score. Across the held-out evaluation set, comprising approximately 1.23 million annotated cells, performance was highest for epithelial cells, which achieved a precision of 0.81, recall of 0.88, and F1 score of 0.85 (Figure 2A). Lymphocytes were also robustly detected (precision, 0.74; recall, 0.80; F1, 0.77). Performance varied across cell types, reflecting differences in abundance, morphology, and histological context. Fibroblasts showed intermediate performance (F1, 0.63), followed by vascular-associated cells (F1, 0.58) and plasma cells (F1, 0.52). Smooth-muscle cells, macrophages, and neutrophils were more challenging to resolve, with F1 scores of 0.47, 0.42, and 0.39, respectively. The confusion matrix also showed strong class-specific performance for epithelial cells and lymphocytes, whereas morphologically overlapping populations exhibited increased misclassification among related cell types (Figure 2B). For example, smooth-muscle cells were commonly assigned to fibroblast or vascular classes, while plasma cells were most frequently misclassified as lymphocytes.

**Figure 2.**
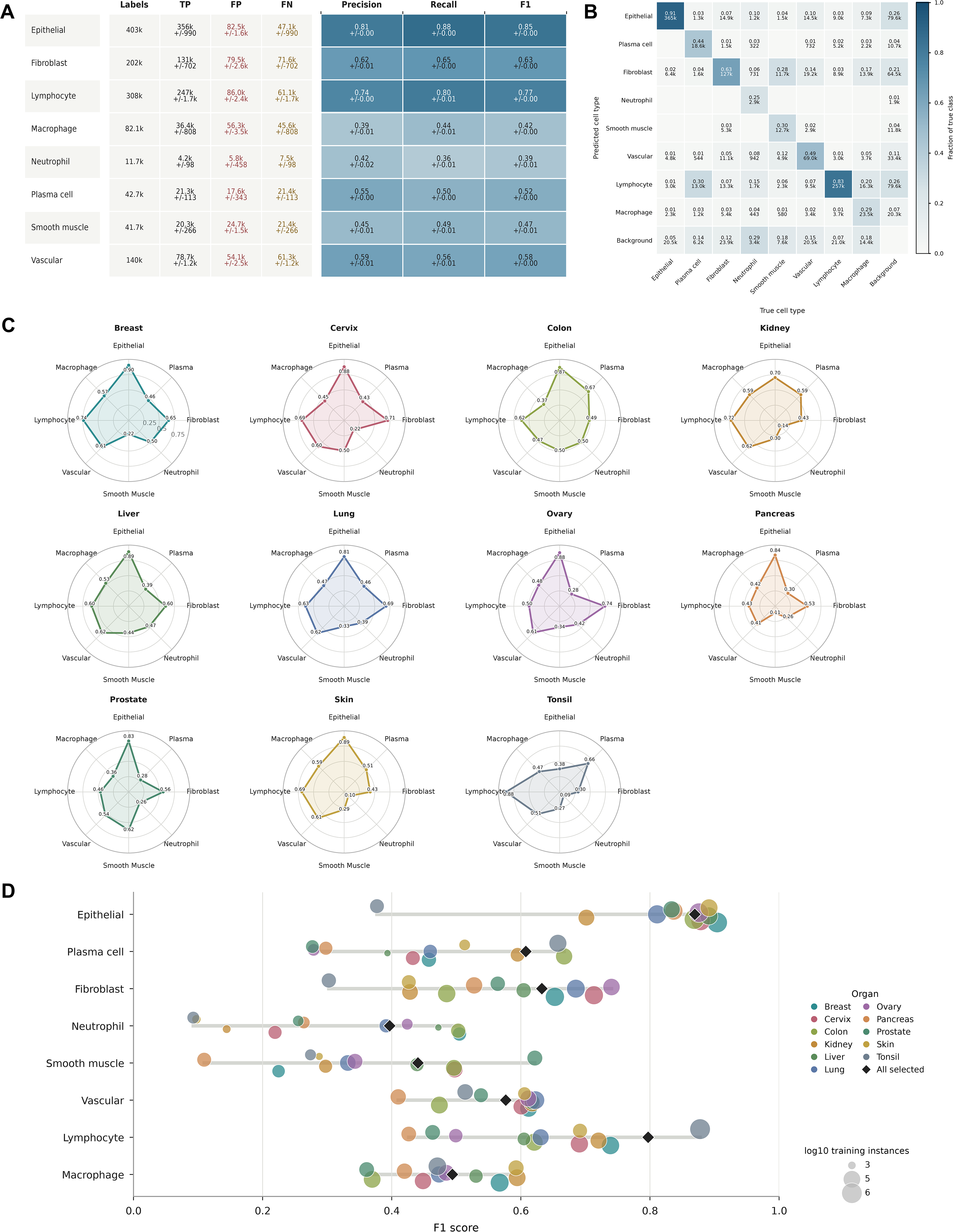
Performance of the HELIOS-trained model across cell types and organs. **(A)** Cell detection and classification performance on the held-out HELIOS evaluation set. The table reports the number of labelled cells, true positives (TP), false positives (FP), false negatives (FN), precision, recall, and F1 score for each of the eight cell classes. Performance values are shown as mean ± standard deviation across five cross-validation folds. **(B)** Normalised confusion matrix showing predicted versus true cell classes, including background detections. Values indicate the fraction of each true class assigned to the corresponding predicted class; cell counts are shown beneath the proportions. **(C)** Organ-specific F1 scores for each cell class across the different organs represented in the held-out evaluation data. **(D)** Relationship between organ- and class-specific F1 score and the number of corresponding training instances. Each colour represents one organ, with circle size proportional to the log10-transformed number of training instances. Black diamonds indicate the aggregate F1 score obtained using all organs.

We further assessed whether model performance was consistent across organs. Organ-specific analysis showed that epithelial and lymphocyte classification remained relatively robust across multiple tissue types, whereas stromal and myeloid-lineage populations displayed greater variability (Figure 2C). Cell-type- and organ-resolved F1 scores further revealed that classification performance was influenced by the number of available training instances, with highly represented classes generally showing more stable performance across tissue contexts (Figure 2D).

### HELIOS-trained model shows strong cross-dataset benchmarking performance

To externally evaluate the performance of the HELIOS-trained model, we benchmarked it against models trained on existing nuclear detection and classification datasets, including MoNuSAC^18^, NuCLS^19^ and Lizard^20^, across each held-out test dataset using harmonised cell classes (Figure 3A). As expected, models generally performed best on their corresponding in-domain test sets. Notably, the HELIOS-trained model showed the strongest cross-dataset performance, achieving F1 scores of 0.66 on Lizard, 0.78 on MoNuSAC, and 0.59 on NuCLS. By comparison, models trained on individual external datasets showed larger performance losses when transferred to other cohorts.

**Figure 3.**
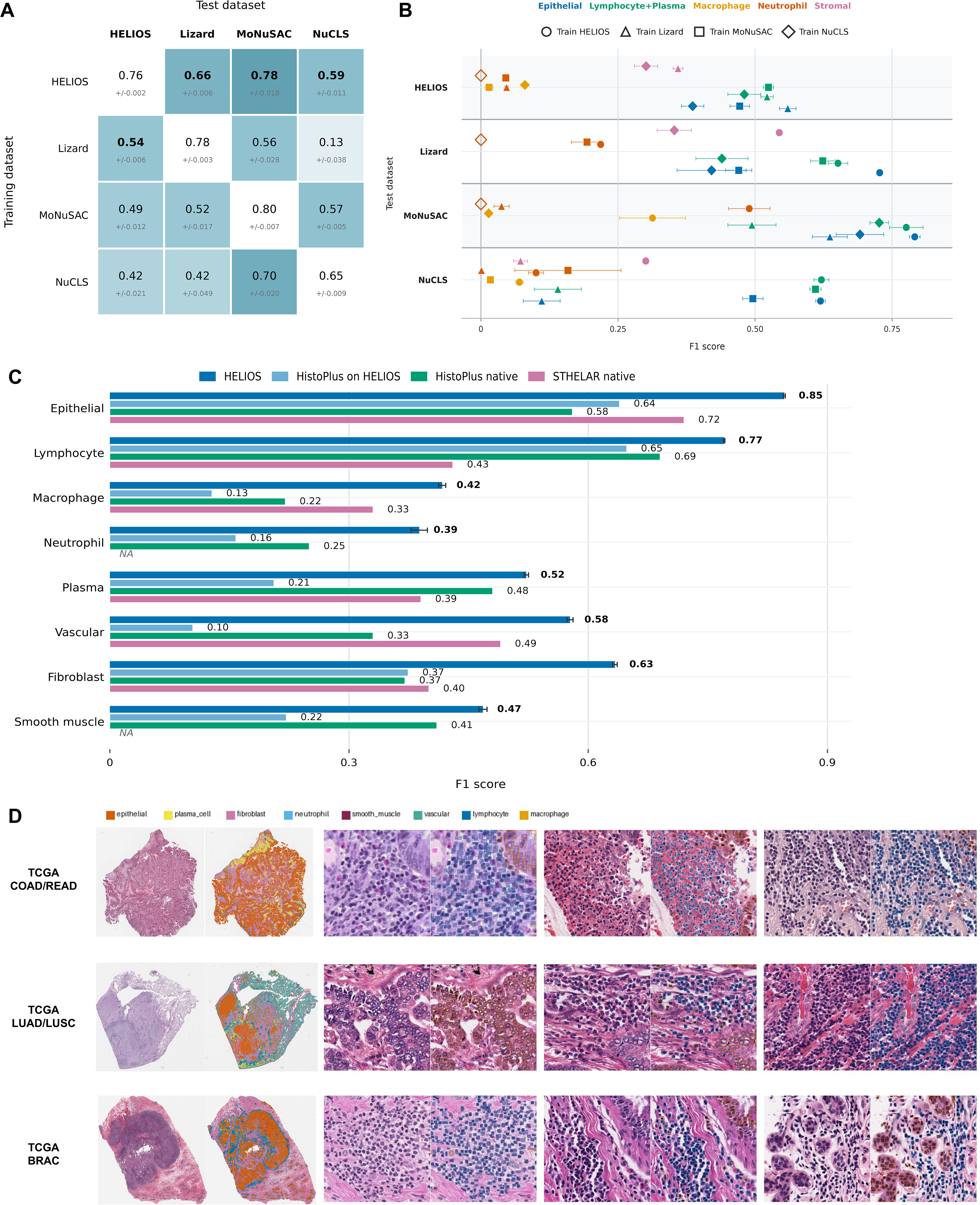
Cross-dataset benchmarking and external evaluation of the HELIOS-trained cell-detection model. **(A)** Cross-dataset comparison of models trained on HELIOS, Lizard, MoNuSAC, or NuCLS and evaluated on each held-out test dataset after harmonisation of cell-class definitions. Cells report the mean overall F1 score ± standard deviation; diagonal entries represent in-domain evaluation, whereas off-diagonal entries measure cross-dataset generalisability. Bold values indicate the highest out-of-domain performance for each test dataset. **(B)** Cell-class-resolved cross-dataset performance. Colours denote cell classes, and marker shapes indicate the dataset used for model training. Points and error bars represent mean F1 scores and their variability on each test dataset. **(C)** Comparison of HELIOS with recent pan-tissue cell-analysis frameworks on the held-out HELIOS test set. The native HELIOS model was compared with the HistoPLUS architecture tested on HELIOS, the native HistoPLUS model, and the native STHELAR model. Bars show class-specific F1 scores; error bars indicate variability across evaluation runs. *NA* indicates that the corresponding class was not available from the comparator model. **(D)** Representative whole-slide and high-magnification predictions from external TCGA cohorts.

Class-resolved benchmarking further demonstrated that the HELIOS-trained model retained the highest F1 scores across all cell compartments, including epithelial, lymphocyte/plasma, macrophage, neutrophil, and stromal categories (Figure 3B). In contrast, a substantial degradation in F1 score was observed for models trained on other datasets when evaluated out of domain.

We further benchmarked HELIOS against two recent pan-tissue cell-analysis frameworks (Figure 3C). HistoPLUS^28^ is a pan-cancer model for nuclear detection and classification trained on a curated dataset of 108,722 nuclei spanning 13 cell types, whereas STHELAR^29^ is a spatial transcriptomics-informed resource comprising more than 11 million cells, whose annotations were used to fine-tune CellViT for multiclass nuclear segmentation and classification. Because the HistoPLUS test dataset was unavailable and some source samples overlapped between STHELAR and HELIOS, a fully independent head-to-head evaluation was not possible. We therefore evaluated the native HistoPLUS and STHELAR models on the held-out HELIOS test set and additionally retrained the HistoPLUS architecture using the HELIOS train set. HELIOS achieved higher F1 score across every evaluated class, including epithelial cells (0.78), lymphocytes (0.75), macrophages (0.40), neutrophils (0.37), plasma cells (0.75), vascular-associated cells (0.58), fibroblasts (0.63), and smooth-muscle cells (0.63). The gains were particularly pronounced for plasma, vascular, fibroblast, smooth-muscle, and myeloid populations, which are less consistently represented in existing histopathology datasets. Representative image overlays from three external cohorts obtained from TCGA confirmed that the HELIOS-trained model could localise and classify diverse cell populations in complex histological regions, including tumour-rich, immune-infiltrated, and stromal compartments (Figure 3D).

### HELIOS reveals conserved and cancer-specific prognostic immune states

We next assessed whether HELIOS-derived immune-cell measurements captured clinically relevant tumour microenvironment states across colorectal, breast, and lung cancers. The HELIOS model was applied to diagnostic H&E-stained whole-slide images from TCGA-COAD/READ, TCGA-BRCA, and TCGA-LUAD/LUSC to quantify tumour-infiltrating lymphocytes (TILs), tumour-associated macrophages (TAMs), and tumour-associated neutrophils (TANs) within tumour-associated regions. Progression-free survival (PFS) and disease-specific survival (DSS) were then compared between high- and low-density subgroups. Representative images illustrated marked variation between immune-cold tumours, characterised by sparse immune-cell infiltration, and immune-hot tumours containing dense lymphocytic and myeloid infiltrates (Figure 4A), together with the distributions of immune-cell densities across tumour types (Figure 4B).

**Figure 4.**
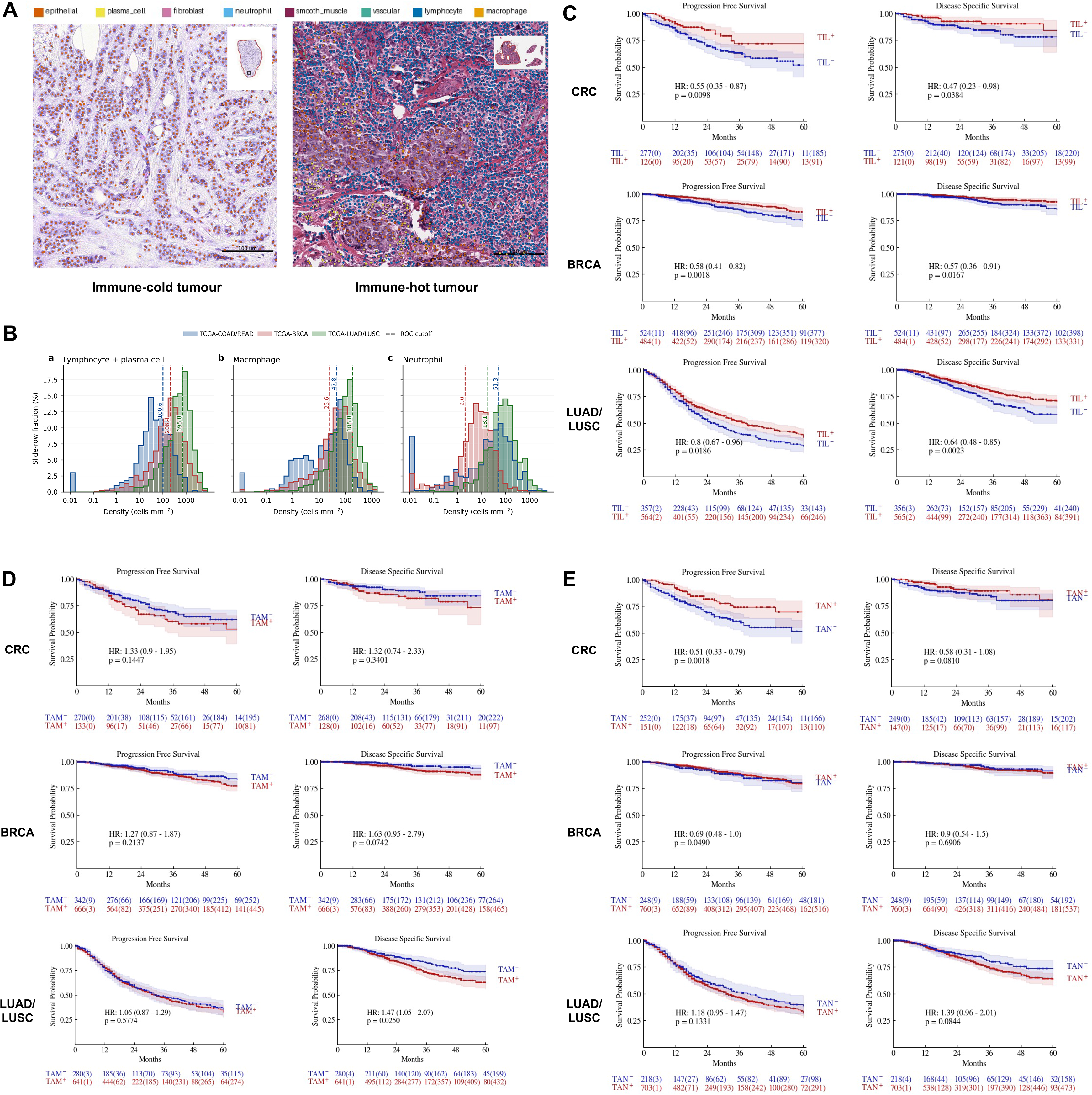
HELIOS-derived immune-cell profiles stratify clinical outcomes across colorectal, breast, and lung cancers. **(A)** Representative HELIOS predictions illustrating immune-cold and immune-hot tumour microenvironments in H&E-stained whole-slide images. Detection overlays show epithelial cells, lymphocytes, plasma cells, macrophages, neutrophils, fibroblasts, vascular cells, and smooth-muscle cells within tumour-associated regions. **(B)** Distributions of tumour-infiltrating lymphocyte (TIL) density, tumour-associated macrophage (TAM), and tumour-associated neutrophil (TAN) densities in TCGA-COAD/READ, TCGA-BRCA, and TCGA-LUAD/LUSC. Dashed vertical lines indicate cohort-specific cut-offs used to define high- and low-density groups. **(C–E)** Kaplan–Meier analyses of progression-free survival and disease-specific survival according to **(C)** TIL density, **(D)** TAM density, and **(E)** TAN density. Within each panel, rows correspond to colorectal, breast, and lung cancer cohorts, respectively. Hazard ratios with 95% confidence intervals and log-rank *p* values are shown. Shaded regions indicate 95% confidence intervals, and numbers at risk are displayed beneath each plot.

Across all three cancer groups, high TIL density was consistently associated with favourable clinical outcome (Figure 4C). In colorectal cancer, TIL-high patients were associated with prolonged progression-free survival (PFS; HR 0.55, 95% CI 0.35–0.87; p = 0.0098) and disease-specific survival (DSS; HR 0.47, 95% CI 0.23–0.98; p = 0.0384). Similarly, in breast cancer, high TIL density was associated with improved PFS (HR 0.58, 95% CI 0.41–0.82; p = 0.0018) and DSS (HR 0.57, 95% CI 0.36–0.91; p = 0.0167). TIL density also stratified outcome in the combined LUAD/LUSC cohort, with TIL-high patients showing longer PFS (HR 0.80, 95% CI 0.67–0.96; p = 0.0186) and DSS (HR 0.64, 95% CI 0.48–0.85; p = 0.0023).

In contrast, associations between TAM density and outcome were generally adverse (Figure 4D). In colorectal cancer, TAM-high patients showed directionally shorter PFS and DSS, although neither association reached statistical significance (PFS: HR 1.33, 95% CI 0.90–1.95; p = 0.1447; DSS: HR 1.32, 95% CI 0.74–2.33; p = 0.3401). Similar trends were observed in breast cancer, including a stronger association with shorter DSS that approached statistical significance (PFS: HR 1.27, 95% CI 0.87–1.87; p = 0.2137; DSS: HR 1.63, 95% CI 0.95–2.79; p = 0.0742). In LUAD/LUSC, TAM density was not associated with PFS but was significantly associated with shorter DSS (HR 1.47, 95% CI 1.05–2.07; p = 0.0250).

TAN density showed cancer-specific prognostic effects (Figure 4E). In colorectal cancer, high TAN were associated with substantially longer PFS (HR 0.51, 95% CI 0.33–0.79; p = 0.0018), with a similar but non-significant association for DSS (HR 0.58, 95% CI 0.31–1.08; p = 0.0810). High TAN density was also weakly associated with favourable PFS in breast cancer (HR 0.69, 95% CI 0.48–1.00; p = 0.0490), but did not stratify DSS. Conversely, TAN-high lung cancers showed directionally poorer outcomes, including shorter PFS (HR 1.18, 95% CI 0.95–1.47; p = 0.1331) and DSS (HR 1.39, 95% CI 0.96–2.01; p = 0.0844).

### HELIOS links histological immune states to molecular tumour subtypes

We further investigated whether HELIOS-derived immune phenotypes reflected molecularly defined tumour states (Figure 5). In colorectal cancer, microsatellite instability-high (MSI) tumours contained significantly higher TAM (p = 0.0003), TAN (p < 0.0001), and overall immune-cell densities (p = 0.0004) than microsatellite-stable tumours (MSS), whereas TIL density did not differ significantly (p = 0.340; Figure 5A). Within MSS colorectal tumours, high TIL density was associated with longer PFS (HR 0.56, 95% CI 0.34–0.92; p = 0.0212), as was high TAN density (HR 0.50, 95% CI 0.31–0.80; p = 0.0040; Figure 5B). The corresponding DSS associations were directionally favourable but did not reach statistical significance.

**Figure 5.**
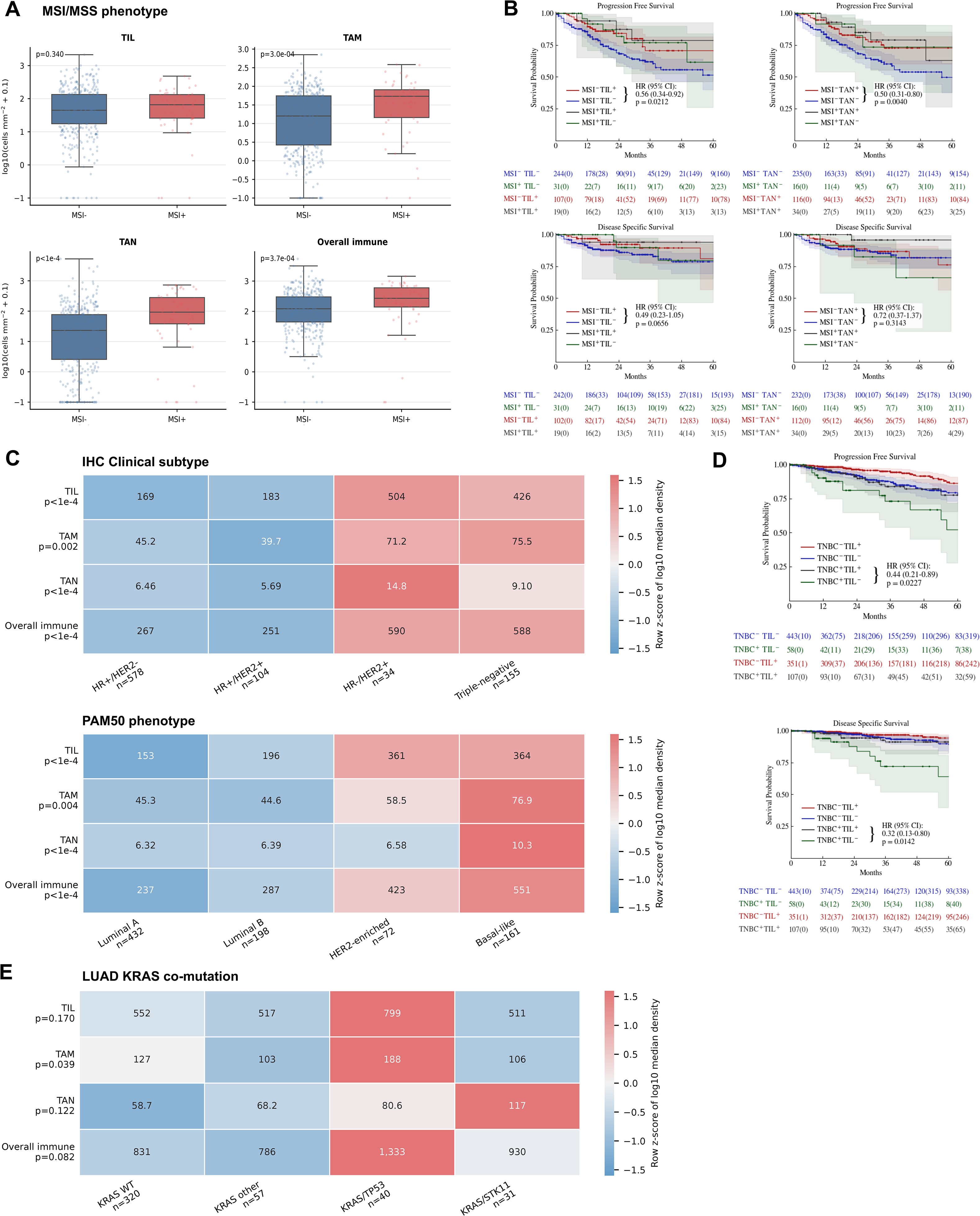
HELIOS-derived immune profiles reflect molecularly defined tumour states in colorectal, breast, and lung cancers. **(A)** Comparison of tumour-infiltrating lymphocyte (TIL), tumour-associated macrophage (TAM), tumour-associated neutrophil (TAN), and overall immune-cell densities between microsatellite-stable (MSI−) and microsatellite instability-high (MSI+) colorectal cancers in TCGA-COAD/READ. Cell densities were log10-transformed, and *p* values were calculated using two-sided Mann–Whitney U tests. **(B)** Kaplan–Meier analyses of progression-free survival and disease-specific survival jointly stratified by MSI status and TIL or TAN density. Hazard ratios, 95% confidence intervals, and log-rank *p* values are shown for the indicated within-subgroup comparisons; shaded regions represent 95% confidence intervals, and numbers at risk are displayed below each plot. **(C)** Immune-cell composition across clinically defined breast cancer receptor groups and PAM50 molecular subtypes in TCGA-BRCA. Colours represent row-wise z-scores of log10-transformed median cell density, while values within each tile show the untransformed median density in cells mm⁻². Overall differences were assessed using Kruskal-Wallis tests. **(D)** Kaplan–Meier analyses of progression-free and disease-specific survival jointly stratified by triple-negative breast cancer status and TIL density. **(E)** TIL, TAM, TAN, and overall immune-cell densities across molecularly defined lung adenocarcinoma groups in TCGA-LUAD: KRAS wild type, other KRAS-mutant tumours, KRAS/TP53-mutant tumours, and KRAS/STK11-mutant tumours.

Breast cancers also displayed marked subtype-specific immune landscapes. Among clinically defined receptor groups, HR-negative/HER2-positive and triple-negative breast cancer (TNBC) showed substantially higher TIL and overall immune-cell densities than HR-positive tumours (both p < 0.0001; Figure 5C). TAM density also differed between groups (p = 0.002), with higher values in HR-negative/HER2-positive and triple-negative disease, while TAN density was highest in HR-negative/HER2-positive tumours (p < 0.0001). Consistent patterns were observed across PAM50 subtypes (Figure 5C). HER2-enriched and basal-like tumours were lymphocyte-rich relative to luminal tumours, whereas basal-like cancers showed the highest TAM and overall immune-cell densities. TAN density was also elevated in basal-like disease. Despite their high overall immune content, TIL density further reveals prognostic value within TNBC. TIL-high tumours in this subgroup were associated with longer PFS (HR 0.44, 95% CI 0.21–0.89; p = 0.0227) and DSS (HR 0.32, 95% CI 0.13–0.80; p = 0.0142; Figure 5D).

Finally, we examined HELIOS-derived immune composition across molecularly defined KRAS groups in LUAD (Figure 5E). Global differences in TIL, TAM, TAN, and overall immune-cell density did not reach statistical significance, although exploratory patterns were apparent. KRAS/TP53-mutant tumours showed numerically higher TIL, TAM, and overall immune-cell densities, whereas KRAS/STK11-mutant tumours showed the highest median TAN density.

### HELIOS identifies cancer-associated fibroblast-rich stromal states associated with adverse outcomes

We next evaluated whether HELIOS could quantify fibroblast-to-epithelial ratio (FER) and identify clinically relevant stromal phenotypes from routine H&E images. Whole-slide inference revealed pronounced spatial heterogeneity in fibroblast abundance, ranging from samples with limited fibroblast component to fibroblast-rich samples (Figure 6A).

**Figure 6.**
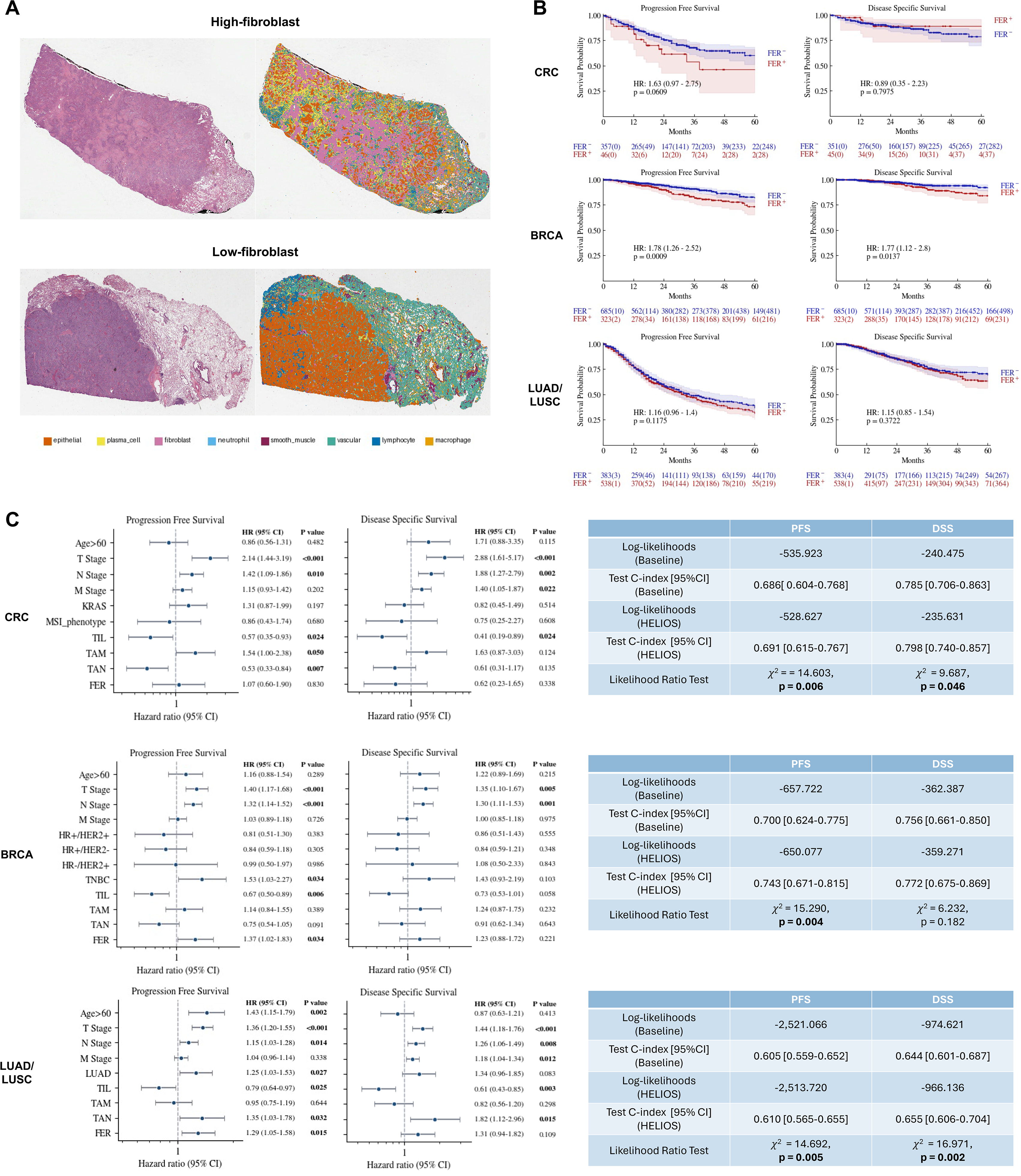
HELIOS identifies prognostically relevant cancer-associated fibroblast-rich stromal states and provides information beyond clinicopathological variables. (A) Representative H&E-stained whole-slide images and corresponding HELIOS prediction overlays illustrating tumours with low and high fibroblast abundance. Fibroblast-to-epithelial ratio (FER) was quantified as the proportion of fibroblasts among epithelial and fibroblast cells within tumour-associated regions. (B) Kaplan–Meier analyses of progression-free survival (PFS) and disease-specific survival (DSS) according to FER-low and FER-high status in TCGA-COAD/READ, TCGA-BRCA, and TCGA-LUAD/LUSC. Hazard ratios with 95% confidence intervals and log-rank *p* values are displayed. Shaded regions indicate 95% confidence intervals, and numbers at risk are shown beneath each plot. (C) Multivariable Cox proportional-hazards analyses of PFS and DSS in colorectal, breast, and lung cancer cohorts. Models incorporated available clinicopathological and molecular variables together with HELIOS-derived tumour-infiltrating lymphocyte (TIL), tumour-associated macrophage (TAM), tumour-associated neutrophil (TAN), and FER measurements. Points indicate hazard ratios, horizontal lines indicate 95% confidence intervals, and statistically significant *p* values are shown in bold. (D) Comparison of baseline clinicopathological Cox models with models additionally incorporating HELIOS-derived cellular features. Tables report model log-likelihoods, test-set C-index with 95% confidence intervals, and likelihood-ratio tests comparing the nested models for PFS and DSS in colorectal, breast, and lung cancers.

A high FER was most strongly associated with adverse outcomes in breast cancer (Figure 6B). Patients with FER-high tumours had significantly shorter progression-free survival (PFS; HR 1.78, 95% CI 1.26–2.52; p = 0.0009) and disease-specific survival (DSS; HR 1.77, 95% CI 1.12–2.80; p = 0.0137). In colorectal cancer, FER-high tumours showed a trend towards shorter PFS (HR 1.63, 95% CI 0.97–2.75; p = 0.0609), whereas no association was observed for DSS (HR 0.89, 95% CI 0.35–2.23; p = 0.7975). In the combined LUAD/LUSC cohort, high FER was directionally associated with shorter PFS (HR 1.16, 95% CI 0.96–1.40; p = 0.1175) and DSS (HR 1.15, 95% CI 0.85–1.54; p = 0.3722), although neither association reached statistical significance.

### HELIOS-derived cellular features provide prognostic information beyond clinicopathological variables

To assess whether HELIOS-derived immune and stromal features provided prognostic information independent of established clinicopathological factors, multivariable Cox models were constructed separately for colorectal, breast, and lung cancers, incorporating age, tumour stage, cohort-specific molecular or histological variables, and HELIOS-derived TIL, TAM, TAN densities and FER (Figure 6C).

In colorectal cancer, TIL density remained independently associated with longer progression-free survival (PFS; HR 0.58, 95% CI 0.35–0.96; p = 0.033) and disease-specific survival (DSS; HR 0.42, 95% CI 0.19–0.93; p = 0.032) after adjustment for stage, KRAS status, and MSI phenotype. High TAN density was also independently associated with longer PFS (HR 0.59, 95% CI 0.35–0.99; p = 0.044), whereas high TAM density was associated with shorter PFS (HR 1.58, 95% CI 1.02–2.45; p = 0.040). FER was not independently associated with either endpoint.

In breast cancer, high TIL density remained independently associated with longer PFS (HR 0.67, 95% CI 0.50–0.89; p = 0.006), whereas high CAF density was associated with shorter PFS (HR 1.37, 95% CI 1.02–1.83; p = 0.034), after adjustment for age, stage, and clinically defined subgroups. TIL density showed a similar favourable association with DSS that approached statistical significance (HR 0.73, 95% CI 0.53–1.01; p = 0.058), whereas the other HELIOS-derived features were not independently associated with DSS.

In the LUAD/LUSC cohort, TIL density was independently associated with longer PFS (HR 0.79, 95% CI 0.64–0.97; p = 0.025) and DSS (HR 0.61, 95% CI 0.43–0.85; p = 0.003). By contrast, TAN density was associated with shorter PFS (HR 1.35, 95% CI 1.03–1.78; p = 0.032) and DSS (HR 1.82, 95% CI 1.12–2.96; p = 0.015). FER was also independently associated with shorter PFS (HR 1.29, 95% CI 1.05–1.58; p = 0.015), but not DSS.

We further evaluated whether adding HELIOS-derived cellular features improved model likelihood and test C-index beyond clinicopathological variables alone (Figure 5C). In colorectal cancer, inclusion of TIL, TAM, TAN densities and FER significantly improved model fit for both PFS (likelihood-ratio test, χ² = 14.603; p = 0.006) and DSS (χ² = 9.687; p = 0.046). The test C-index increased from 0.686 to 0.691 for PFS and from 0.785 to 0.798 for DSS. In breast cancer, HELIOS-derived features significantly improved the PFS model (χ² = 15.290; p = 0.004), with the test C-index increasing from 0.700 to 0.743. The corresponding improvement for DSS was not statistically significant (χ² = 6.232; p = 0.182), although the test C-index increased from 0.756 to 0.772. In LUAD/LUSC, the addition of HELIOS features improved model fit for both PFS (χ² = 14.692; p = 0.005) and DSS (χ² = 16.971; p = 0.002), with test C-indices increasing from 0.605 to 0.610 and from 0.644 to 0.655, respectively.

## Discussion

HELIOS was developed to test the hypothesis that clinically meaningful cellular phenotypes can be captured from routine histology by AI at scale, and spatial omics can provide sufficiently scalable and biologically grounded supervision. In this study, the findings support this premise at three levels. First, molecularly assigned cell identities could be transferred to matched H&E morphology across multiple organs and spatial profiling technologies. Second, models trained using these annotations retained performance across external histopathology datasets with different tissue compositions and annotation schemes. Third, the resulting cellular measurements captured molecularly and clinically relevant tumour microenvironment states in independent patient cohorts. HELIOS therefore extends cell identification using spatial omics beyond the tissues directly profiled: molecular information derived from a comparatively limited set of deeply characterised samples can be propagated to substantially larger collections of routine H&E slides.

This strategy is conceptually distinct from predicting visual-omics from H&E-stained images. Although recent visual-omics models demonstrate that selected transcriptional programmes can be inferred from tissue morphology^21,22^, many molecular states are transient, context-dependent, or only weakly encoded in H&E appearance. HELIOS instead learned from spatial transcriptomics to define cellular identity during model development and restricts the downstream prediction task to phenotypes that are sufficiently represented by morphology. This approach provides a practical balance between biological resolution and clinical scalability: spatial omics supplies the molecular ground truth, whereas routine histology provides the substrate for broad deployment.

Classification performance nevertheless varied across cellular compartments, revealing both the strengths and current limits of morphology-based cell recognition. Epithelial cells and lymphocytes were detected most accurately, likely because they have comparatively distinctive morphological features and were well represented in the training data. By contrast, myeloid and stromal populations showed greater confusion, consistent with their overlapping nuclear and cytoplasmic appearances and the biological continuity between related cell states. Similar limitations were observed in the comparator models, suggesting that this challenge is not specific to HELIOS but reflects a broader constraint of cell classification from H&E morphology alone. Despite this, HELIOS showed the strongest overall performance across the evaluated datasets and cell classes, including comparatively underrepresented immune and stromal populations. These findings indicate that the scale, organ diversity, and spatial omics-informed supervision of HELIOS improve generalisability, while also highlighting the need for hierarchical classification, uncertainty-aware prediction, and more extensive representation of rare and phenotypically overlapping cell types.

The prognostic importance of immune-cell density and spatial localisation has been established across multiple malignancies^30–33^. The clinical analyses using TCGA cohorts showed that HELIOS recovered a conserved lymphocyte-associated tumour state across colorectal, breast, and lung cancers using a single model applied to routine H&E images. Importantly, TIL density remained associated with outcome after adjustment for stage and tumour-specific molecular variables, indicating that HELIOS-derived immune profiling captures information beyond established clinicopathological risk factors.

The clinical implications of myeloid infiltration were strongly dependent on tumour context. This was particularly evident for TANs, which were associated with a significant favourable outcome in colorectal cancer but an adverse outcome in lung cancer. In colorectal cancer, several studies have associated densities of neutrophils with improved survival^34–37^. In this study, the enrichment of TANs in MSI-high colorectal cancers and their favourable prognostic association may therefore reflect an organised, immune-active microenvironment.

Conversely, in lung cancer, neutrophil-rich microenvironments may arise from chronic inflammatory programmes ^38,39^. Direct mechanistic evidence is particularly strong for STK11/LKB1 loss. In KRAS-driven lung adenocarcinoma models, STK11 deficiency induces neutrophil recruitment and pro-inflammatory cytokine production while reducing intratumoural T-cell activity^40^. KRAS/STK11 tumours in our analysis (Figure 5E) also showed the highest median TAN density but only modest overall immune infiltration. This pattern may represent a neutrophil-dominant, lymphocyte-attenuated state that supports immune evasion and tumour progression.

More broadly, studies have demonstrated that tumour-associated neutrophils comprise diverse and plastic states rather than a uniform lineage. These include inflammatory, cytotoxic, immunosuppressive, angiogenic, and antigen-presenting programmes, with their relative abundance shaped by tumour type and local signalling^41,42^. The opposing prognostic associations observed here may therefore reflect differences in neutrophil state or cellular neighbourhood rather than neutrophil abundance alone.

The molecular subtype analyses recapitulated established immune landscapes across colorectal, breast, and lung cancers (Figure 5), demonstrating that routine H&E contains informative morphological features to recover clinically relevant biological states. This raises the possibility that HELIOS could serve as a rapid, low-cost screening tool to identify tumours with characteristic immune phenotypes or molecular subtypes before comprehensive genomic profiling, helping prioritise patients for downstream molecular testing. The analysis also revealed substantial heterogeneity among tumours within the same molecular group. MSI-high colorectal cancers were broadly immune enriched, consistent with their elevated neoantigen burden and characteristic lymphocytic infiltration^43,44^. Nevertheless, in our cohort, TIL and TAN densities retained prognostic value within microsatellite-stable disease. HELIOS may therefore help identify immune-active states among the majority of colorectal cancers that lack the MSI-high phenotype. Similarly, triple-negative breast cancers were among the most immune-rich breast tumours. This observation is consistent with pooled clinical evidence showing that TIL abundance is strongly prognostic in early triple-negative breast cancer. Importantly, TIL density still separated patients with markedly different outcomes^45^. Given the aggressive clinical course and limited subtype-defining therapeutic targets of triple-negative disease, reproducible H&E-based quantification could provide a scalable means of resolving prognostic heterogeneity within this high-risk population^46^.

Beyond immune-cell infiltration, HELIOS-derived fibroblast abundance was most strongly associated with adverse outcomes in breast cancer and remained independently associated with progression in both breast and lung cancers. This finding is consistent with breast cancer studies showing that enrichment of activated fibroblast populations is associated with recurrence, metastasis, and poorer survival^47,48^. Spatial single-cell profiling in non-small-cell lung cancer showed that tumour-like and matrix-associated cancer-associated fibroblast (CAF) phenotypes were linked to poor survival, whereas inflammatory and interferon-responsive CAF states were associated with more favourable outcomes^49^. These findings support the clinical relevance of the stromal compartment but also emphasise that total FER is a coarse measure of a heterogeneous cell population. Integrating molecularly defined fibroblast states may distinguish fibroblast populations that promote immune exclusion and tumour progression from those with tumour-restraining functions.

The multivariable analyses provide evidence that HELIOS-derived measurements contain information beyond established clinicopathological variables across cancer types. Adding cellular features consistently improved model likelihood and C-index, and likelihood-ratio results indicate that the tumour microenvironment contributes independent prognostic information. These findings indicate that molecular classification alone does not fully capture the cellular organisation of tumours or the associated variation in clinical risk. HELIOS-derived phenotypes may therefore provide additional stratification, particularly for patients who lack targetable genomic alterations or established predictive biomarkers.

The broader translational value of HELIOS is its ability to extract multiple cellular phenotypes from material already embedded in diagnostic workflows. Once trained, a histology-based model can be applied retrospectively to archived clinical-trial material and population-scale cohorts without further tissue consumption. This creates an opportunity to test cellular hypotheses in datasets that would be infeasible to profile comprehensively using spatial assays. Importantly, HELIOS generates spatially resolved cell maps that can be directly reviewed by pathologists, providing a visually interpretable link between model-derived measurements and the underlying tissue morphology. The same development framework could also be adapted to rare cancers or treatment-specific cohorts by adding targeted spatial omics samples to a shared multi-organ reference. To support clinical translation and reproducibility, we developed a web-based platform (https://octopath.ai) that enables users to upload H&E-stained whole-slide images and run HELIOS inference. The platform is currently undergoing evaluation across multiple clinical centres.

### Limitations of the study

First, HELIOS resolves eight major cell populations rather than the full spectrum of molecularly and functionally distinct cell states. Macrophage, neutrophil, fibroblast, and lymphocyte subsets can have divergent biological and clinical effects despite overlapping appearances on H&E. Moreover, the lower classification performance observed for several myeloid and stromal populations indicates that their abundance is estimated less precisely than that of epithelial cells and lymphocytes. Larger and more diverse spatial-omics datasets, together with hierarchical and uncertainty-aware modelling, may enable more refined inference of clinically relevant cellular states.

Second, although spatial-omics-derived labels provide biologically grounded supervision, they are not error-free. Residual uncertainty in cell-type assignment may arise from image co-registration, cell segmentation and differences among spatial profiling platforms. Confidence-based filtering and benchmarking against manual annotations may reduce these sources of noise, while filtering low-confidence cells may result in incompletely annotated training images, which can be suboptimal for object-detection frameworks such as YOLOv10. Future work should therefore evaluate training strategies that account explicitly for partially labelled data, as well as alternative instance-segmentation and cell-classification architectures.

Third, the prognostic analyses were retrospective and conducted primarily in TCGA cohorts assembled across multiple institutions, with heterogeneity in clinical tissue preparation and image quality. Some molecularly defined subgroup analyses also included relatively small numbers of patients and should therefore be regarded as exploratory. Although HELIOS-derived features improved multivariable model fit, these associations do not establish causality or treatment-predictive utility. Independent validation in prospectively collected and uniformly treated cohorts, particularly clinical trials of immunotherapy, will be required before clinical implementation.

Finally, the current clinical analyses primarily quantified global cell abundance and did not fully capture spatial organisation or cell-cell interactions. The clinical effect of a cellular population is likely to depend on its location and neighbourhood. Features such as immune exclusion, tumour-fibroblast niches, vascular proximity, and multicellular neighbourhoods may therefore provide more mechanistically informative biomarkers.

In conclusion, we developed HELIOS, a biology-grounded multi-cancer atlas for nuclei detection and classification from routine H&E-stained whole-slide images. The AI model trained using HELIOS demonstrated robust performance across internal and external benchmarking datasets. The recovery of established immune patterns across colorectal, breast, and lung cancers, together with associations between HELIOS-derived cellular phenotypes and patient survival, indicates that HELIOS can extract clinically relevant information about both tumour biology and prognosis beyond established molecular and genomic markers from routine H&E morphology. By transferring cellular knowledge learned from deeply profiled spatial-omics specimens to large collections of routine H&E slides, HELIOS provides a foundation for interpretable tumour-microenvironment profiling and patient stratification at clinical scale.

## Data Availability

All data produced in the present work are contained in the manuscript

## Acknowledgment

This project is funded by UK Research and Innovation (MR/T040785/1). KB was funded by the Swiss National Science Foundation (P500PM_217647/1). MAH is supported by the National Institute for Health and Care Research University College London Hospitals Biomedical Research Centre.

## Methods

### Data Source

#### Spatial transcriptomics data

HELIOS was constructed by integrating 60 publicly available formalin-fixed paraffin-embedded (FFPE) spatial transcriptomics samples, comprising 6 normal and 54 tumour tissues from 14 human organs, including breast, colorectum, lung, cervix, lymph node, ovary, pancreas, liver, kidney, prostate, brain, heart, skin, and tonsil. The spatial transcriptomics data were generated using four complementary platforms: 10x Genomics Xenium In Situ, Xenium Prime, Visium HD, and Atera Whole Transcriptome Analysis (WTA). For each sample, spatial transcriptomics outputs, cell segmentation contours, matched H&E-stained whole-slide images, and associated metadata were downloaded from the original data repositories. Repository URLs for all samples included in this study are provided in Supplementary Table 1.

#### Single-cell RNA-sequencing data

Single-cell RNA sequencing datasets used as reference atlases for cell type annotation were obtained from the CellXGene Database^50^ and the DISCO Single-Cell Omics Database^51^. Healthy tissue reference datasets were collected from CellXGene, whereas tumour reference datasets were obtained from CellXGene and supplemented with tumour-specific datasets from DISCO to improve representation across cancer types. To ensure compatibility across spatial transcriptomics platforms, only genes present in the corresponding spatial transcriptomics gene panels were retained for downstream analyses.

#### Datasets for benchmarking cell detection and classification

To comprehensively evaluate the generalizability of HELIOS, we benchmarked the proposed models on three publicly available datasets that represent diverse tissue types, annotation strategies, and cell taxonomies (Table 1).

**Table 1.** Tissue types and cell classes in the external datasets.

| Dataset | Tissue types | Annotated nuclei | Cell classes |
| --- | --- | --- | --- |
| MoNuSAC | Breast, kidney, lung, prostate | 46,909 | Epithelial, lymphocyte, macrophage, neutrophil |
| NuCLS | Breast | 59,485 | Tumour, fibroblast, lymphocyte, mitotic figure, vascular, plasma, apoptotic, macrophage, neutrophil, myoepithelial, normal epithelial, eosinophil |
| Lizard | Colorectal | 495,179 | Epithelial, lymphocyte, plasma, neutrophil, eosinophil, connective tissue |

MoNuSAC^18^ is a multi-organ nucleus segmentation and classification benchmark comprising H&E image patches from 71 patients across four organs (breast, kidney, lung, and prostate). The dataset contains 46,909 manually annotated nuclei belonging to four major cell categories and provides instance-level segmentation masks together with cell-type labels.

NuCLS^19^ is a breast cancer nuclear annotation dataset derived from TCGA whole-slide images. It contains over 220,000 manually annotated nuclei with localisation, segmentation, and fine-grained cell-type labels. The dataset was developed through a collaborative annotation workflow involving pathologists, pathology trainees, and crowdsourcing with expert adjudication, providing high-quality annotations for computational pathology algorithm development.

Lizard^20^ is one of the largest publicly available colorectal nucleus datasets containing 495,179 annotated nuclei from H&E image patches. The dataset provides instance segmentation masks and six-class nucleus classification labels generated through an automated annotation pipeline followed by extensive pathologist refinement.

#### Datasets for clinical interpretation

To evaluate the clinical and biological relevance of HELIOS-derived phenotypes, we applied the trained HELIOS model to diagnostic H&E-stained whole-slide images from The Cancer Genome Atlas (TCGA) and analysed matched clinicopathological variables and survival endpoints. Three major tumour groups were selected to reflect organs represented in both HELIOS and external clinical cohorts: colorectal cancer, breast cancer, and lung cancer.

For colorectal cancer, TCGA colon adenocarcinoma and rectum adenocarcinoma cohorts were combined and analysed as TCGA-COAD/READ. For breast cancer, TCGA breast invasive carcinoma was analysed as TCGA-BRCA. For lung cancer, the TCGA lung adenocarcinoma and lung squamous cell carcinoma cohorts were analysed separately and jointly as the TCGA-LUAD/LUSC cohort.

#### Cell annotation pipeline

We developed a gene-panel-aware cell annotation pipeline to assign cell identities to spatial transcriptomics-derived cells and generate molecularly informed annotations for matched H&E-stained images. The pipeline consisted of three main stages: construction of organ- and panel-matched single-cell references, platform-specific definition of spatial cell objects, and transcriptomic label transfer from the matched reference to each spatial transcriptomics sample. The workflow was applied independently to each sample to preserve sample-specific gene-panel composition, organ context, and tissue morphology.

#### Construction of organ- and panel-matched single-cell references

Six spatial transcriptomics samples had paired single-cell RNA-sequencing data, which were used directly as matched references for cell annotation. For the remaining samples, we constructed sample-specific single-cell RNA-sequencing references according to the organ type and gene panel used for each spatial transcriptomics assay. For each sample, only genes shared between the spatial transcriptomics dataset and the candidate single-cell reference were retained for downstream analysis. This ensured that label transfer was performed using genes directly measurable in the corresponding spatial transcriptomics platform, reducing bias introduced by differences in gene coverage across assays.

Each constructed reference was initialised using CELLxGENE-derived single-cell data, comprising an organ-specific tissue component and a shared immune-cell component. The tissue component included both normal and tumour-associated cell populations, whereas the shared immune component primarily represented normal immune-cell populations. Reference cells were mapped to a common broad cell-type vocabulary covering T cells, B cells, NK cells, monocytes, macrophages, dendritic cells, plasma cells, mast cells, neutrophils, innate lymphoid cells, epithelial cells, endothelial cells, fibroblasts, stromal cells, mesenchymal cells, pericytes, and smooth muscle cells. The raw labels annotated by the pipeline were then grouped into the final eight classes used for cell detection and classification in H&E images (Table 2). Cells that are not identifiable from H&E-stained morphology were removed from the final dataset.

**Table 2.** Label mapping.

| **Final label in HELIOS** | **Raw Labels in the cell annotation pipeline** |
| --- | --- |
| Epithelial | Epithelial |
| Fibroblast | Fibroblast, Mesenchymal, Stromal |
| Lymphocyte | B, T, NK, Innate Lymphoid |
| Plasma | Plasma |
| Macrophage | Macrophage |
| Vascular | Pericyte, Endothelial |
| Smooth Muscle | Smooth Muscle |
| Neutrophil | Neutrophil |

To reduce cell-type imbalance, each broad cell type was down-sampled to 10,000 cells where sufficient cells were available. Datasets with insufficient cellular representation (less than 100 cells) were excluded from reference construction to avoid unstable or poorly supported reference profiles.

For tumour-profile samples, the CELLxGENE reference was supplemented with tumour-enriched single-cell references from the DISCO Single-Cell Omics Database, where suitable datasets were available. DISCO datasets were filtered according to organ type, cancer or disease term, tumour-like sample type, cell-type confidence, and overlap with the corresponding spatial transcriptomics gene panel. Retained DISCO cells were mapped to the same broad cell-type vocabulary and balanced using the same cell-type capping strategy before integration with the CELLxGENE-derived reference.

The reference source, organ type, and number of genes for each spatial transcriptomics sample are provided in Supplementary Table 2.

#### Platform-specific definition of spatial cell objects

For Xenium and Atera samples, the default cell contours provided by the spatial transcriptomics platform were used as the spatial cell objects for label transfer. The corresponding cell-by-gene expression matrices were loaded together with cell-level metadata and spatial coordinates. Platform-derived cell boundaries were retained during label transfer so that each predicted cell identity could be assigned directly to the corresponding segmented cell object on the matched H&E image.

For Visium HD samples, cell-level objects were inferred before label transfer because the platform provides high-resolution binned expression measurements rather than default single-cell contours. For each sample, the 2 μm binned gene-expression matrix was used to define the measured gene set, and a sample-specific panel file was generated from the detected gene-expression features. Organ-specific references were first constructed using the union of genes detected across Visium HD samples from the same organ and then filtered to the sample-specific gene set before label transfer.

To generate cell-level expression profiles for Visium HD, the matched H&E image was used to initialise image-guided cell segmentation with Cellpose-SAM^52^. These image-derived initial contours were then used with ProSeg^53^ to infer cell boundaries and assign local gene-expression signal to individual cells. The resulting ProSeg-derived cell objects, consisting of cell-level expression profiles and corresponding polygonal boundaries, were used as the query cells for downstream label transfer. This approach enabled Visium HD samples to be processed within the same reference-matched label-transfer framework as Xenium and Atera samples, while accounting for the absence of platform-provided single-cell contours.

#### Transcriptomic label transfer

Cell identities were transferred from the matched single-cell reference to each spatial transcriptomics sample using a gene-panel-aware, PCA-based label transfer strategy^54^. For each sample, the reference and query expression matrices were matched single-cell reference and the spatial transcriptomics assay *G*. The reference harmonised by gene symbol and restricted to the genes shared between the and query matrices were represented as:

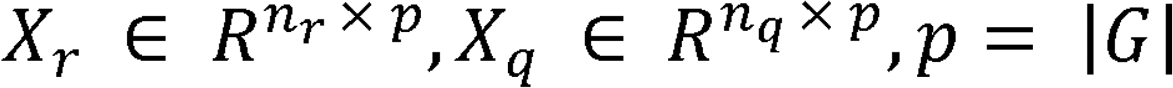

where *n_r_* and *n_q_* are the numbers of reference and query cells, respectively, and *p* is the number of shared genes. Both matrices were normalised and log-transformed before dimensionality reduction.

Principal component analysis was fitted on the reference expression matrix, and query cells were projected into the same reference-defined embedding space:

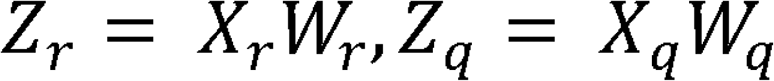

where *w_r_* denotes the PCA loading matrix learned from the reference data, *z_r_* and *z_q_* denote the low-dimensional embeddings of the reference and query cells, respectively.

For each query cell, the *k* nearest reference cells were identified in the shared embedding space using cosine distance. The distance between query cell *i* and reference cell *j* was calculated as:

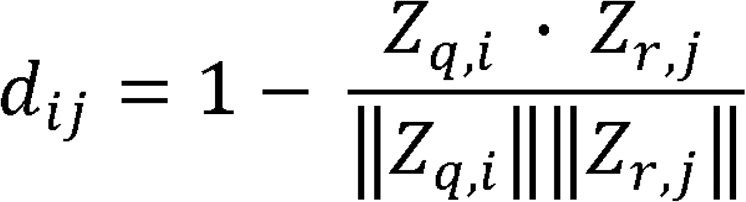

Let *N_k_(i)* denote the set of *k* nearest reference neighbours for query cell *i*, and let *d_i,k_* denote the distance to the *k_th_* nearest neighbour. Each neighbour was assigned a distance-normalised score:

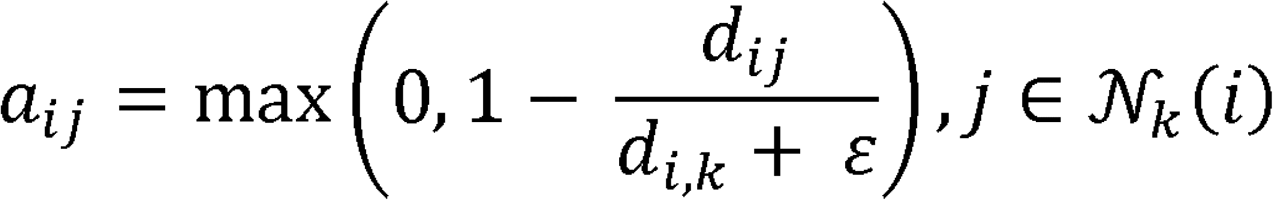

Where ε is a small constant used to avoid division by zero. These scores were then smoothed using a Gaussian kernel and normalised across the nearest-neighbour set:

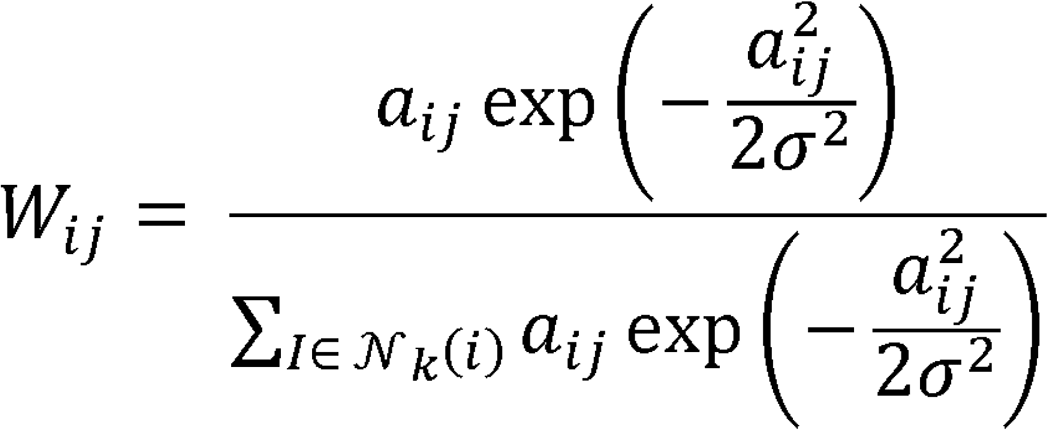

where *w_ij_* represents the final contribution of reference cell *j* to query cell *i*, and σ controls the width of the Gaussian weighting kernel. This weighting scheme assigns greater influence to nearby reference cells while reducing the contribution of distant neighbours within the local neighbourhood.

For each candidate cell type *c*, a label-transfer score was calculated by summing the weights of neighbouring reference cells assigned to that class:

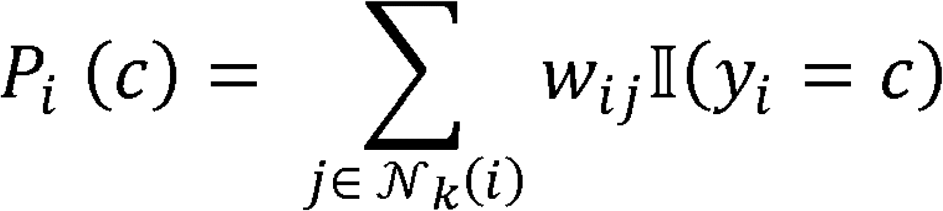

where *Y_i_* is the known label of reference cell *j* and *u*(.) is the indicator function. The predicted identity of query cell *i* was assigned as the class with the highest label-transfer score, and the corresponding prediction score was retained as the maximum class score:

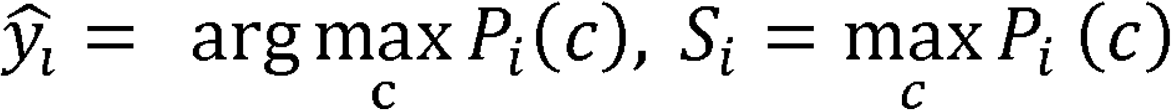

#### Cell contour refinement using Cellpose-SAM

Following transcriptomic label transfer, cell contours were refined using Cellpose-SAM on the matched H&E-stained images to improve the morphological accuracy of the final annotations. For Xenium and Atera samples, platform-provided cell contours were used during label transfer, ensuring that transcriptomic cell identities were assigned to the original spatial transcriptomics-derived cell objects. These transferred labels were then post-processed against H&E-derived Cellpose-SAM contours.

For each sample, Cellpose-SAM was applied within the tissue region covered by the spatial transcriptomics data. Tiled inference was used to segment cells across the corresponding H&E image, generating image-derived candidate cell contours. Transferred transcriptomic labels were then matched to Cellpose-SAM-derived contours according to spatial overlap. A transferred cell label was retained if its original spatial transcriptomics-derived contour intersected an H&E-derived Cellpose-SAM contour. For retained cells, the predicted cell identity and associated confidence score from transcriptomic label transfer were preserved, while the final cell boundary was replaced by the matched Cellpose-SAM contour. Labels without a corresponding H&E-derived contour were excluded from the refined annotation set and used for quality-control assessment.

### Quality control of the cell annotation pipeline

To support quality control, additional neighbourhood-based confidence metrics were calculated. The neighbourhood distance spread and normalised spread were defined as:

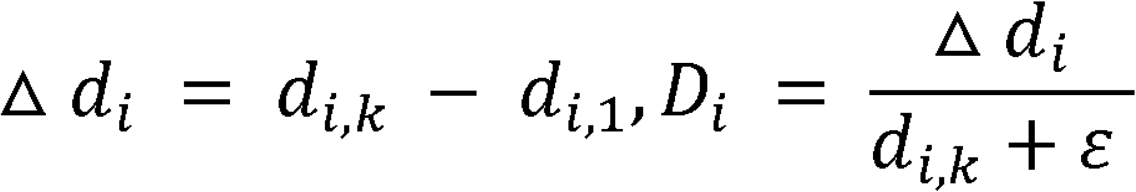

where *d_i,1_* is the distance to the nearest reference neighbour. A combined confidence score was then calculated as:

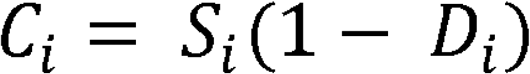

The combined confidence score integrates the label-transfer prediction score with local neighbourhood compactness; higher values therefore indicate query cells with both strong class-assignment support and a compact local reference neighbourhood.

To evaluate the accuracy of the cell annotation pipeline, we compared pipeline-derived cell labels with pathologist annotations in a manually reviewed subset of cells. Cells were randomly selected across four confidence-score intervals: 0-0.25, 0.25-0.5, 0.5-0.75, and 0.75-1.0. Performance was assessed for each cell class using precision, recall, F1-score, and class support. Across 611 manually annotated cells, the pipeline achieved a weighted-average precision of 0.739, recall of 0.785, and F1-score of 0.755. Among cells with confidence scores greater than 0.5, performance increased substantially, with a weighted-average precision of 0.857, recall of 0.842, and F1-score of 0.842, indicating that high-confidence annotations showed strong agreement with pathologist review. Given that 90% of transferred labels across the full dataset had confidence scores greater than 0.5, the resulting annotations support a high-fidelity cell-labelled dataset for downstream model training and evaluation.

### AI model development and evaluation

#### Model training

We trained a YOLOv10-m model for cell detection and classification using the HELIOS dataset. Image patches of 640×640 pixels were extracted from annotated H&E-stained whole-slide images, together with the corresponding cell bounding boxes and class labels. The model was trained for 200 epochs using 8 NVIDIA A100 GPUs. The training batch size was 48, and model optimisation was performed using the AdamW optimiser with an initial learning rate of 0.01.

Colour augmentation was applied to the training data to reduce the impact of staining variability and improve model robustness across samples, tissues, and data sources. Stain separation was applied to deconvolved RGB channels into haematoxylin and eosin channels. Stain concentration perturbation strategy was applied to introduce realistic stain variation, in which perturbation factors were uniformly sampled with σ = 0.14 and applied to the deconvolved H&E channels before reconstructing the augmented RGB images. Spatial augmentations were also applied during training, with configuration parameters listed in Supplementary Table 3.

#### Internal evaluation

Internal model performance was evaluated using five-fold cross-validation. In each fold, the dataset was split into training, validation, and held-out test subsets at a ratio of 80:10:10. The model was trained on the training subset, hyperparameters were selected using the validation subset, and final performance was reported on the held-out test subset. This process was repeated across five folds, and performance metrics were summarised across folds. Model performance was assessed using precision, recall, and F1-score at the cell level. Per-class metrics were reported to assess performance across individual cell populations, and weighted-average metrics were used to summarise overall model performance.

#### External evaluation

To assess generalisability beyond HELIOS, the trained model was externally evaluated on three publicly available nucleus segmentation and classification benchmarks: Lizard, MoNuSAC, and NuCLS, which include key cell groups shared in HELIOS and tissue types across colorectal, breast, lung, kidney, and prostate. Cell labels from each external dataset were harmonised to the closest corresponding HELIOS cell classes. The trained HELIOS model was applied without further fine-tuning to each external dataset. Performance was assessed using the same classification metrics as in the internal evaluation.

#### Biomarker Quantification

##### Tumour segmentation

Tumour-associated regions for colorectal cancers were identified on H&E-stained whole-slide images using an EfficientNet^55^ trained on the NCT-CRC-HE-100K^56^ dataset. The network was trained for 100 epochs on a single NVIDIA GeForce RTX 3090 GPU with a batch size of 256 using the AdamW optimiser and an initial learning rate of 1 × 10. Data augmentation included random horizontal flipping (probability = 0.5) and stain-based colour augmentation to improve robustness to inter-slide staining variability. The same colour augmentation strategy used for YOLOv10 training was applied to the EfficientNet model.

Tumour-associated regions for breast and lung were identified using the ICGI automatic tumour segmentation model^57^. This model generates a tumour probability map for each whole-slide image, which is subsequently thresholded to produce a binary tumour segmentation mask. The model was developed and validated as a pan-cancer tumour segmentation approach for FFPE H&E whole-slide images across multiple carcinoma types.

##### Cell density

Cell densities were reported as the number of detected cells per mm² within the tumour-associated regions. Lymphocytes and plasma cells were combined into a single density measure, while macrophages and neutrophils were quantified separately:

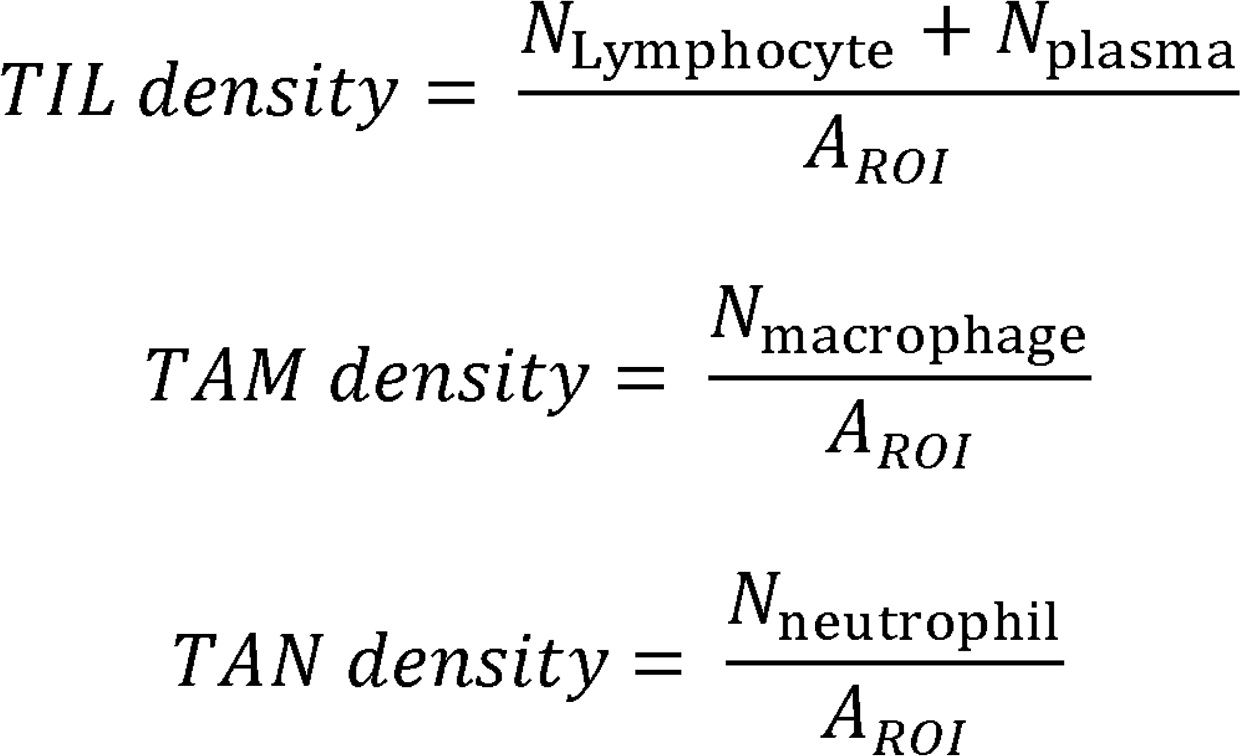

where N denotes the number of detected cells of the corresponding class within the tumour-associated region, and *A_ROI_* denotes the segmented tumour-associated area in mm².

The FER was defined as the proportion of fibroblasts among epithelial and fibroblast cells in the tumour-associated area:

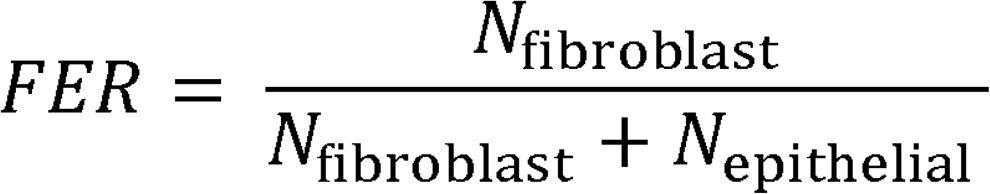

For survival and subgroup analyses, biomarker cut-off values were optimised using ten-fold cross-validation with five-year disease progression as the endpoint. Within each fold, candidate thresholds were evaluated for their ability to distinguish patients with and without disease progression within five years, and the threshold maximising Youden’s J index was selected and applied in downstream analyses.

##### Statistics

For comparisons of HELIOS-derived cell densities across molecular or clinicopathological subgroups, two-group comparisons were performed using the Mann-Whitney U test, and comparisons across more than two groups were performed using the Kruskal-Wallis test. Categorical variables were compared using the chi-square test or Fisher’s exact test, as appropriate. Associations between continuous biomarkers were assessed using Spearman’s rank correlation. For survival analyses, progression-free survival and disease-specific survival were analysed using Kaplan-Meier curves and compared between biomarker-defined groups using the log-rank test. Hazard ratios and 95% confidence intervals were estimated using Cox proportional hazards regression.

Multivariable Cox regression models were used to assess whether HELIOS-derived features provided prognostic information independent of clinicopathological variables. Models were constructed separately for colorectal, breast, and lung cancer cohorts and adjusted for available cohort-relevant variables, including age, tumour stage, and molecular or histological subgroup annotations where available. To evaluate the added value of HELIOS-derived biomarkers, nested Cox models with and without HELIOS features were compared using likelihood-ratio tests. Model discrimination was assessed using the concordance index.

All statistical tests were two-sided, and p<0.05 was considered statistically significant. Subgroup analyses and multi-biomarker comparisons were interpreted as exploratory unless otherwise specified. Complete-case analysis was used for models requiring clinical, molecular, imaging, and survival variables.

## Notes

### Competing Interest Statement

The authors have declared no competing interest.

### Author Declarations

The study used (or will use) ONLY openly available human data that were originally located at: https://portal.gdc.cancer.gov/

